# Rapid, Affordable and Scalable SARS-CoV-2 Detection from Saliva

**DOI:** 10.1101/2021.04.01.21254182

**Authors:** Andrew Hayden, Marcy Kuentzel, Sridar V. Chittur

## Abstract

Here we present an inexpensive, rapid, and robust RT-LAMP based SARS-CoV-2 detection method that is easily scalable, enabling point of care facilities and clinical labs to determine results from patients’ saliva directly in 30 minutes for less than $2 a sample. The method utilizes a novel combination of widely available reagents that can be prepared in bulk, plated and frozen and remain stable until samples are received. This innovation dramatically reduces preparation time, enabling high-throughput automation and testing with time to results (including setup) in less than one hour for 96 patient samples simultaneously when using a 384 well format. By utilizing a dual-reporter (phenol red pH indicator for end-point detection and SYTO-9 fluorescent dye for real-time), the assay also provides internal validation of results and redundancy in the event of an instrument malfunction.

## INTRODUCTION

Since early 2020 the world has been overtaken by the Covid-19 pandemic caused by infections of the Sars-Cov2 coronavirus. There have been many efforts by various groups around the globe to develop molecular and antigen-based assays for detection and surveillance of this infection. Early efforts using RT-qPCR^1,2^ based detection of nasopharyngeal swabs from individuals were quickly hampered by disruption of supply chains for reagents and consumables and insufficient testing sites across the world. These challenges led scientists to look for alternative methods for detection using a wide variety of biosamples that included oral/nasal swabs, mouth gargles^3^, saliva^2,4–6^ and urine^7,8^. The detection methodologies also included non-RT-qPCR methods such as reverse transcription loop isothermal amplification (RT-LAMP^2,9–11^), CRISPR-based^12–14^, reverse transcription recombinase polymerase amplification (RT-RPA)^15^ or reverse transcription recombinase aided amplification (RT-RAA)^16^ with their outputs coupled with fluorescent or colorimetric reporters as well as lateral flow strip platforms to facilitate readout processes. Some of the popular methods still included the requirement to isolate the virus from the sample and this was a cause of concern again due to the extra steps/consumables required. The requirements for expensive instrumentation, reagents and time prevents some of these assays from being widely deployable especially to low resource settings. Here we describe our modifications of the RT-LAMP assay using saliva that is inexpensive, quick and scalable for various resource settings and does not require RNA extraction.

During our protocol development, we noticed that the standard LAMP assay protocol tended to generate false positive reactions (albeit at later time points) and we questioned if inclusion of the loop primers were necessary. Previous reports from other groups using LAMP have suggested that the loop primers could result in reproducibility issues with LAMP. Our analysis suggested that while exclusion of loop primers delayed the reactions, we could detect amplification reliably by omitting just the loop forward (LF) primer from the master mix. This observation resulted in our recipe with just 5 of the 6 commonly used primers for each gene in the LAMP protocol. Our assays included primer sets for the E and the N genes and is called EN* primer mix (Figure 1).

**Figure 1:**
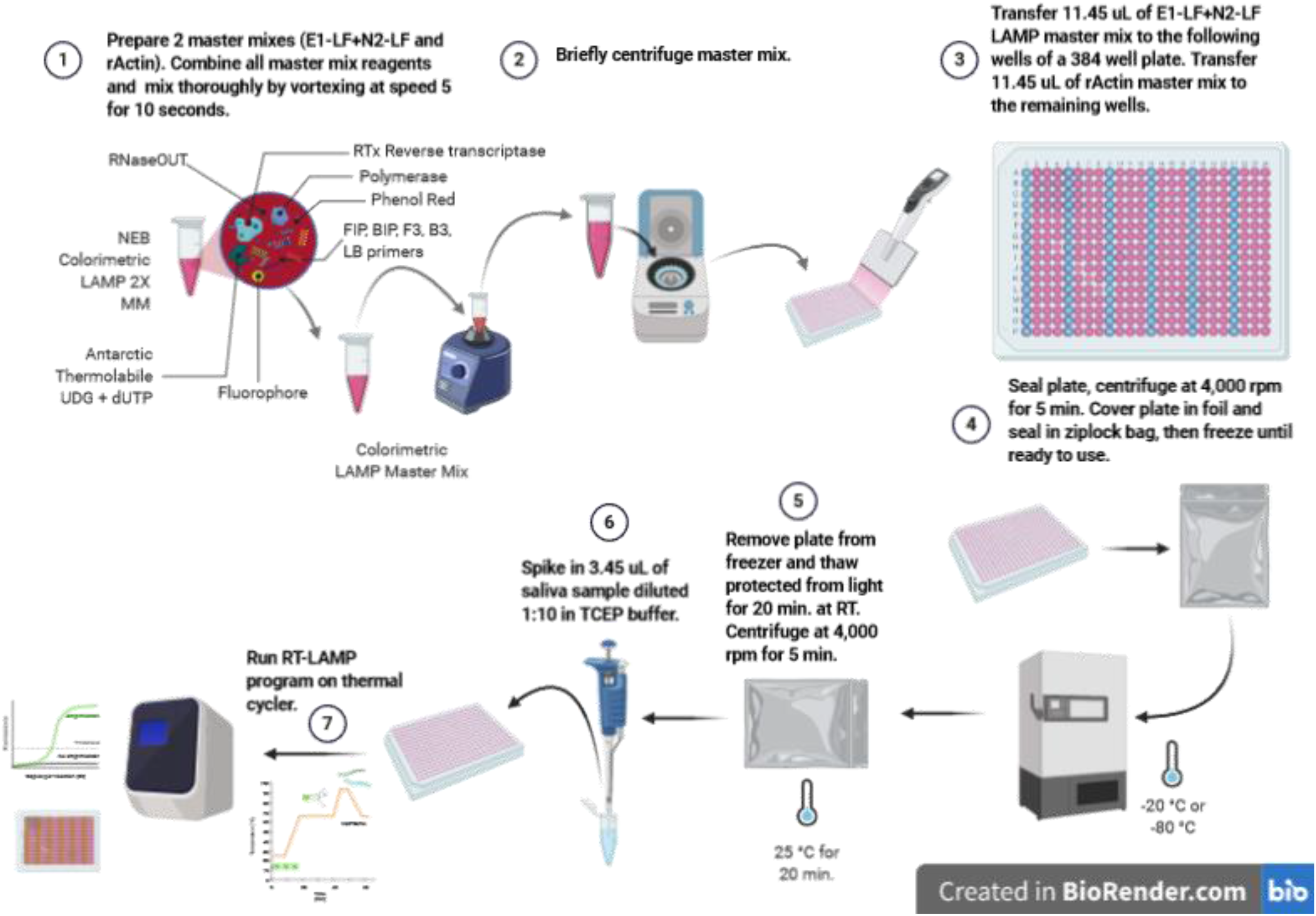
Schematic of dual-assay workflow

## METHODS AND MATERIALS

The assay is a modified version of the colorimetric RT-LAMP protocol developed by Tanner *et al*. at New England Biolabs ^2,4,17–19^, which includes guanidine hydrochloride for enhanced sensitivity and specificity. Further modifications included the addition of Antarctic thermolabile UDG to reduce carryover contamination thereby reducing false-positives, and the inclusion of RNaseOUT recombinant RNase inhibitor or polyvinylsulfonic acid (PVSA) for improved sample stability^20,21^. The saliva sample preparation method is modified from Rabe and Cepko’s protocol^22^.

The following reagents used in assay development were purchased from New England BioLabs: WarmStart^®^ Colorimetric LAMP 2X Master Mix (DNA & RNA) (M1800L), WarmStart^®^ LAMP Kit (DNA & RNA) (E1700L), Antarctic Thermolabile UDG (M0372L), dUTP Solution (N0459S). All primers used for early assay development were standard-desalted oligos synthesized by Integrated DNA Technologies (IDT). Assay optimization was performed using HPLC purified inner primers (FIP and BIP) for SARS-CoV-2 E1 and N2 genes synthesized by LGC Biosearch Technologies. Internal control primers for rActin were from NEB SARS-CoV-2 Rapid Colorimetric LAMP assay kit (Cat. No. E2019S). Guanidine hydrocholoride (GuHCl) (Millipore CAS No. 5010) and tris (2-carboxyethyl) phosphine (TCEP) (Sigma-Aldrich C4706) were generously donated by Dr. Qishan Lin of the RNA Epitranscriptomics & Proteomics Resource, SUNY Albany. Other reagents were purchased from Thermo Scientific and Sigma Aldrich: Invitrogen UltraPure 0.5M EDTA, pH 8.0 (Cat. No. 15575020), Invitrogen RNaseOUT™ Recombinant RNase Inhibitor (Cat. No. 10777019), Invitrogen SUPERase·In™ (Cat. No. AM2696), Sigma Aldrich polyvinylsulfonic acid, sodium salt solution 30% wt. in H2O (Cat. No. 278424) and VWR Nuclease-free Water (not DEPC-Treated) (Cat. No. 02-0201-1000). Positive controls used for assay validation included synthetic control RNA from Twist Bioscience (SARS-CoV-2 Control 1 MT007544.1, Cat. No. 102019) and these BEI Resources: genomic RNA from SARS-CoV-2 isolate USA-CA3/2020 (NR-52507), SARS-CoV-2 isolate USA-WA1/2020 heat-inactivated cell lysate (NR-52286), and SARS-CoV-2 isolate USA-WA1/2020 gamma-irradiated cell lysate (NR-52287).

### Reaction Preparation

All reagents were thawed at room temperature and vortexed gently but thoroughly 10 seconds at medium speed, then quickly spun down and placed (except NEB Colorimetric LAMP 2X Master Mix and NEB 50X Fluorescent dye) on ice until ready to assemble. The 50X fluorescent dye was diluted 1:50 in pure DMSO in an amber 1.5 mL tube or covered with foil. Diluted 1X fluorescent dye was vortexed briefly and spun down and then stored protected from light at room temperature until assembly (dye will precipitate if placed on ice).

### LAMP Master Mix (15 µL Reaction Volume) setup

The RT-LAMP reaction mixes (see supplementary table 1) included two primer mixes (the E1-LF + N2-LF or EN* mix for the COVID-19 test, and the rActin mix as an internal control).

The following reagents were combined in two separate 1.5 mL microcentrifuge tubes in this order:

1. 7.5 µL NEB Colorimetric LAMP 2X Master
2. 0.8 µL 1X Fluorescent Dye
3. 1.5 µL 10X LAMP Primers (EN* / rActin)
4. 0.3 µL Antarctic Thermolabile UDG (2U/µL)
5. 5. 0.1 µL dUTP (100 mM)
6. 0.5 µL GuHCl (1.2 M)
7. 0.75 µL RNaseOUT (**30U/15 µL reaction**) or PVSA **(900 µg/mL)**

Mix thoroughly by vortexing at medium speed for 10 seconds, spin down and protect from light until ready to plate.

Using a single/multichannel pipette 11.45 µL of EN* master mix was aliquoted into columns 2-4, 6-8, 10-12, 14-16, 18-20, and 22-24 of a 384 well optical plate. The rActin (11.45 µL) master mix was aliquoted into columns 1, 5, 9, 13, 17, and 21. After sealing the plate with optical tape, we centrifuged at 4,000 rpm for 5 minutes to remove bubbles and settle contents in wells. This plate was cover with aluminum foil and seal in a zip lock bag and frozen at −20°C until ready to use.

### Saliva Sample Preparation

Upon receipt, saliva was heat-inactivated at 95°C for 5 minutes, then placed on ice immediately for 3-10 minutes to chill. After cooling, the saliva was centrifuged at 5,000 rpm or greater for 5 minutes to pellet debris. The supernatant was removed while carefully avoiding the pelleted material and transferred to a fresh 1.5 mL tube. This supernatant was then diluted 1:5x or 1:10x with 2.5 mM TCEP + 1 mM EDTA buffer prepared as per Rabe & Cepko’s protocol^22^, and pipetted 10X to mix thoroughly, spun down and stored on ice until ready to assay. When saliva samples were unable to be processed immediately, the diluents were frozen at –80 °C for upwards of two weeks.

### Running the Assay

On removal from freezer the plate was thawed at room temperature protected from light 20 minutes before running the assay. Once thawed, the plate seal was removed, and 3.55 µL of positive control was spiked in into wells in columns 2, 6, 10, 14, 18, and 22; 3.55 µL of NTC (water) was spiked into wells in columns 3, 7, 11, 15, 19, and 23; then saliva samples diluted in 2.5 mM TCEP 1mM EDTA pH 8.0 were spiked in into wells in columns 1, 4, 5, 8, 9, 12, 13, 16, 17, 20, 21, and 24. After resealing the plate with optical tape, it was vortexed briefly to mix followed by centrifugation at 4,000 rpm for 5 minutes. The assay was performed on a thermal cycler or real-time qPCR instrument with the following settings:

- 65°C for 30 minutes (data collection every 30 seconds for 80 cycles if using qPCR)
- 4°C for 5 minutes (or melt-curve analysis if using qPCR)

## Results

Initial testing was conducted using NEB WarmStart^®^ Colorimetric LAMP 2X Master Mix (DNA & RNA) (M1800L) and WarmStart^®^ LAMP Kit (DNA & RNA) (E1700L) using N-A and ORF1a primers from Dao Thi *et al*. ^19^, but due to low reproducibility and high false positive rates with these primers, further assay development used E1 and N2 primer sets designed by Zhang *et al*. ^18^ combined with 40 mM guanidine hydrochloride. The E1 and N2 primers with guanidine yielded dramatic improvements in sensitivity and specificity of the assay, demonstrating a limit of detection of 10 copies per reaction using Twist Bioscience SARS-CoV-2 control RNA. However, despite these improvements the assay still suffered sporadic non-specific amplification in no template controls. To eliminate carryover contamination as a potential cause of non-specific amplification, Antarctic thermolabile UDG and dUTP were added to the reaction mixture. These components succeeded in quashing NTC amplification, but amplification in positive control reactions remained inconsistent.

### Primer mix optimization

To address this short-coming, both E1 and N2 primers were analyzed using IDT’s OligoAnalzyer tool to determine whether cross-reactivity such as primer-dimer or secondary structures in any of the primers might lead to inconsistent amplification as Meagher *et al*. suggest ^23^. OligoAnalyzer results revealed that some primer pairs could form exceptionally strong secondary structures even at the LAMP incubation temperature of 65°C. Additionally, it was found the E1-LF primer had a melting-temperature 20°C lower than any other primer in either the E1 or N2 primer sets, suggesting that it could inhibit amplification by out competing the other primers during hybridization. As Khorosheva, E. M., *et al*.^24^ suggest in their research using digital real-time RT-LAMP, LAMP primers must hybridize to the template in a specific order to enable amplification to occur (F3/B3 → FIP/BIP → LF/LB), any other order terminates amplification abruptly. Interestingly, Khorosheva *et al*. also found in their research that although loop primers improve the speed of amplification in LAMP, these primers do not improve the reaction efficiency, such that a RT-LAMP reaction containing a single loop primer can be as efficient as or more so than a reaction containing two loop primers. Ding *et al*.^25^ also found that removing loop primers from a LAMP reaction can improve specificity and sensitivity. Based on the OligoAnalyzer data and the literature, all SARS-CoV-2 LAMP primer combinations were evaluated systematically to determine the optimum sets (see Figure 2). The combined LAMP primer sets E1 and N2 that omitted the LF primers from both sets demonstrated the best specificity, sensitivity and time to positive, so all further tests used this optimum primer combination.

**Figure 2:**
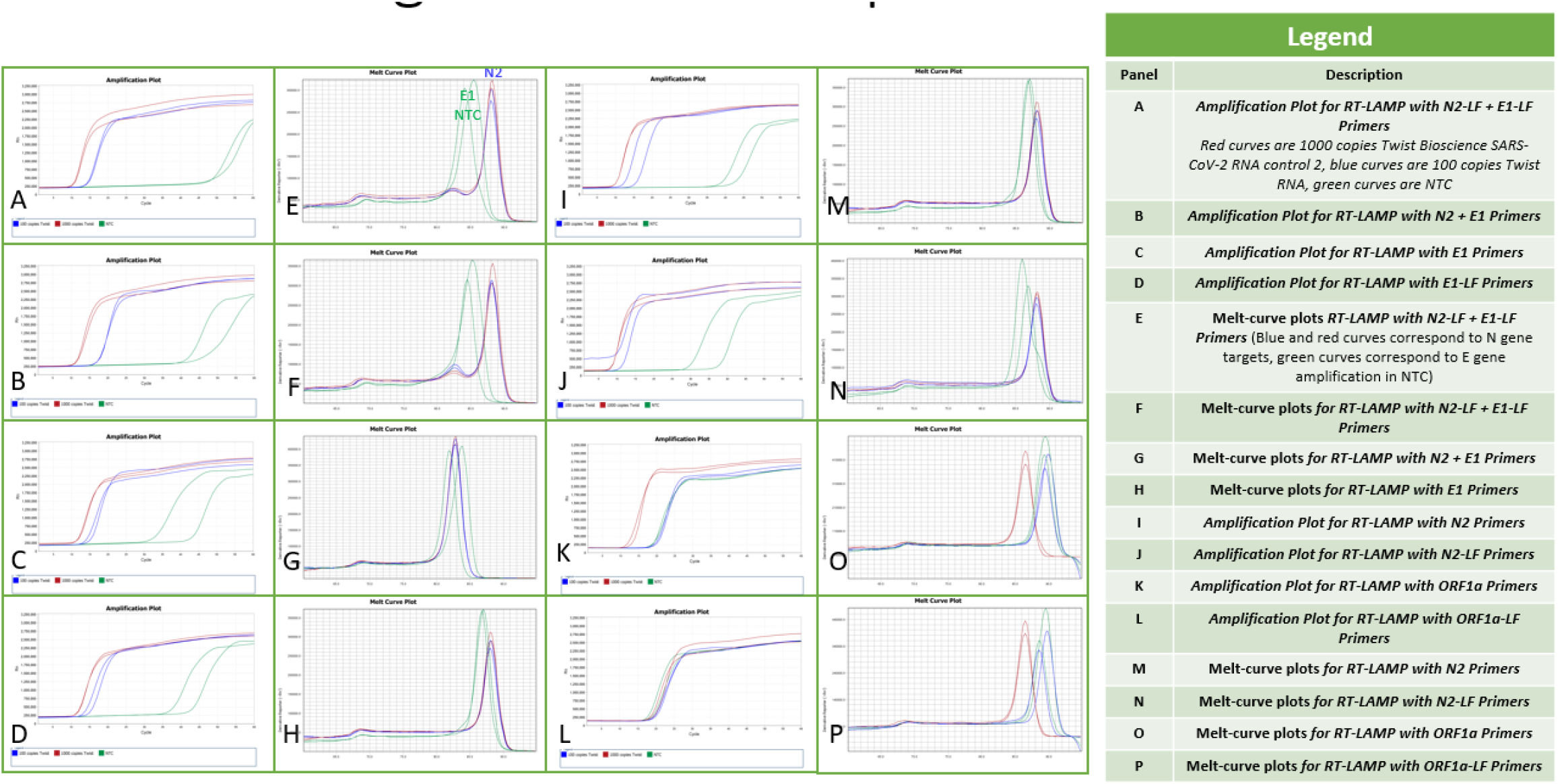
Primer Optiization

### Sample RNA integrity optimization

After primer optimization, sensitivity and specificity testing of direct saliva RT-LAMP was pursued. Initial sensitivity tests using mock positive saliva (saliva that was spiked with either Twist control for Sars-Cov2 or BEI coronavirus cell lysate) gave inconsistent results, sometimes showing no amplification even with high RNA template inputs. These results indicated that active nucleases were present in the heat inactivated saliva samples. To prevent further degradation of RNA that could result in false-negative tests, various RNase inhibitors were tested both in the sample preparation and the RT-LAMP master mix. The following RNase inhibitors were evaluated: NEB RNase inhibitor murine, Invitrogen SUPERase·In, and Invitrogen RNaseOUT Recombinant Ribonuclease Inhibitor. Only RNaseOUT at a final concentration of 30 units per reaction was found to consistently inhibit nuclease activity in direct saliva RT-LAMP at the optimum incubation temperature of 65°C. RNaseOUT performed equally well when used either in the master mix or applied to the saliva sample.

Sensitivity and specificity testing (Figure 3) were conducted using negative saliva samples that were heat inactivated at 95°C for 5 minutes^21^ and diluted 1:10 in TCEP buffer before being spiked with BEI SARS-CoV-2 isolate USA-WA1/2020 heat-inactivated cell lysate (NR-52286) (stock concentration = 375,000 copies/uL) to final concentrations of 1000, and 100 total genome copy equivalents (GCE) per reaction. All reactions were run with 20 replicates. No template controls contained negative saliva diluted 1:10 in TCEP buffer.

Sensitivity and specificity for mock saliva tests containing 1000 copies or 100 copies of SARS-CoV-2 RNA were both 100% with combined E1-LF and N2-LF primer sets.

**Figure 3:**
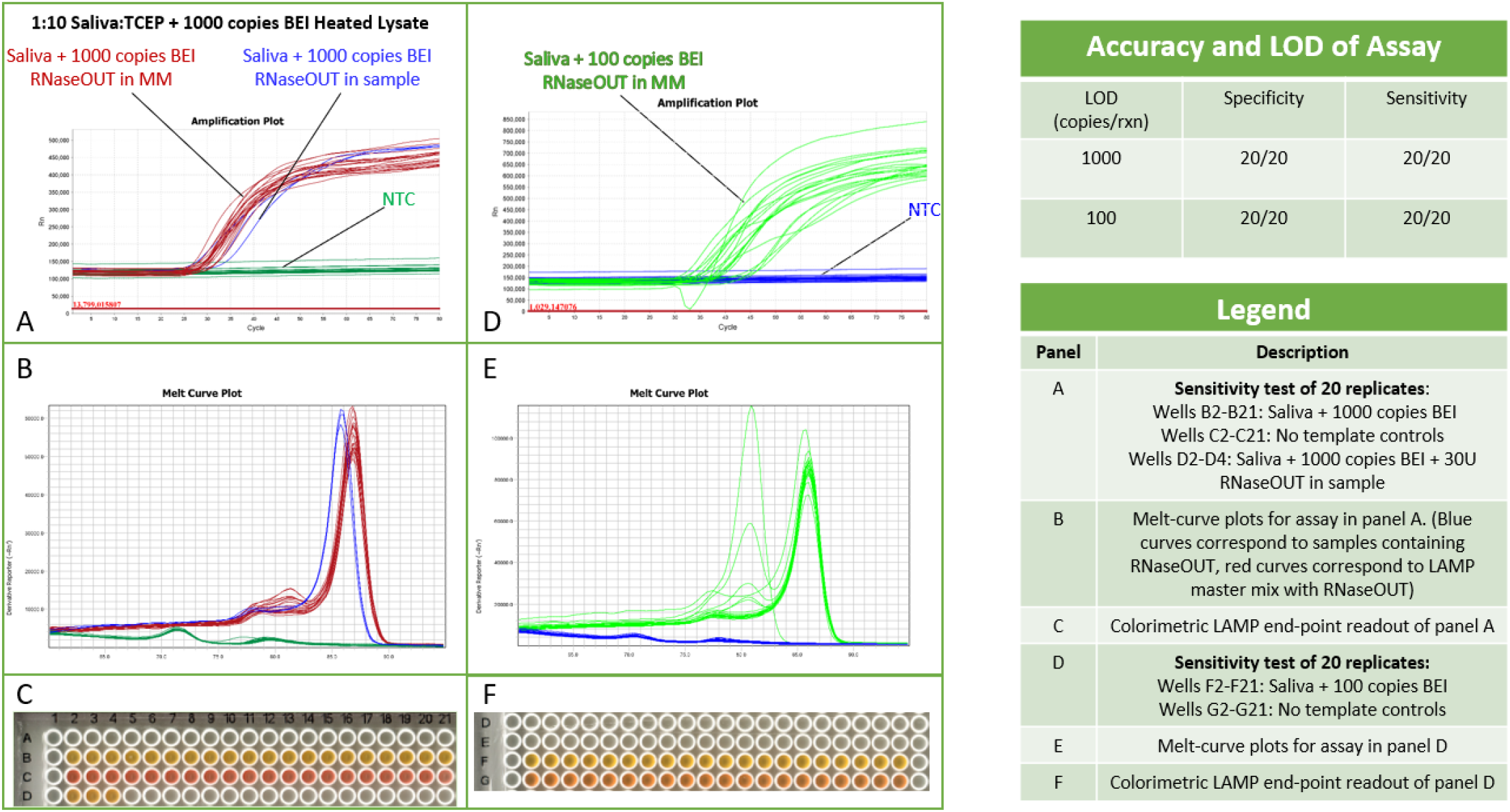
Sensitivity and Specificity testing

### Assay cost reduction

Despite the dramatic improvement in reproducibility of the assay upon inclusion of RNaseOUT, it was felt that the cost of this nuclease inhibitor would be prohibitive especially in low resource settings. To reduce cost and improve accessibility of the assay, chemical nuclease inhibitors were also investigated. A search of the literature^20,21,26^ revealed that the anionic polymer polyvinylsulfonic acid (PVSA) is an exceptionally potent, thermostable, yet affordable RNase inhibitor when used in RT-qPCR, IVT, and RT-LAMP. Earl et al.^20^ demonstrated that PVSA could improve mRNA integrity fivefold when used in IVT at less than 1/1,700^th^ the cost of conventional ribonuclease inhibitors, and Smyrlaki et al.^21^ showed that PVSA at a final concentration of 45 µg/mL dramatically improved SARS-CoV-2 RNA stability in heat inactivated nasopharyngeal swabs in their direct RT-qPCR assay. PVSA also lyses cells as Yu, Racevskis, and Webb^26^ described, thereby improving RNA yield and potentially decreasing the limit of detection for direct detection from patient samples.

Comparative sensitivity and specificity testing was done to evaluate the effectiveness of PVSA against RNaseOUT using the same conditions as previously described. These tests showed that PVSA outperformed RNaseOUT at preserving the integrity of RNA in saliva samples (Figure 4). RT-LAMP master mix containing a final concentration of 45 µg/mL PVSA demonstrated better reproducibility and quicker time to positive results than master mixes containing 30 U/reaction RNaseOUT. There were no false positives or false negatives, demonstrating the assay’s exceptional robustness when testing saliva directly (Figure 5).

**Figure 4:**
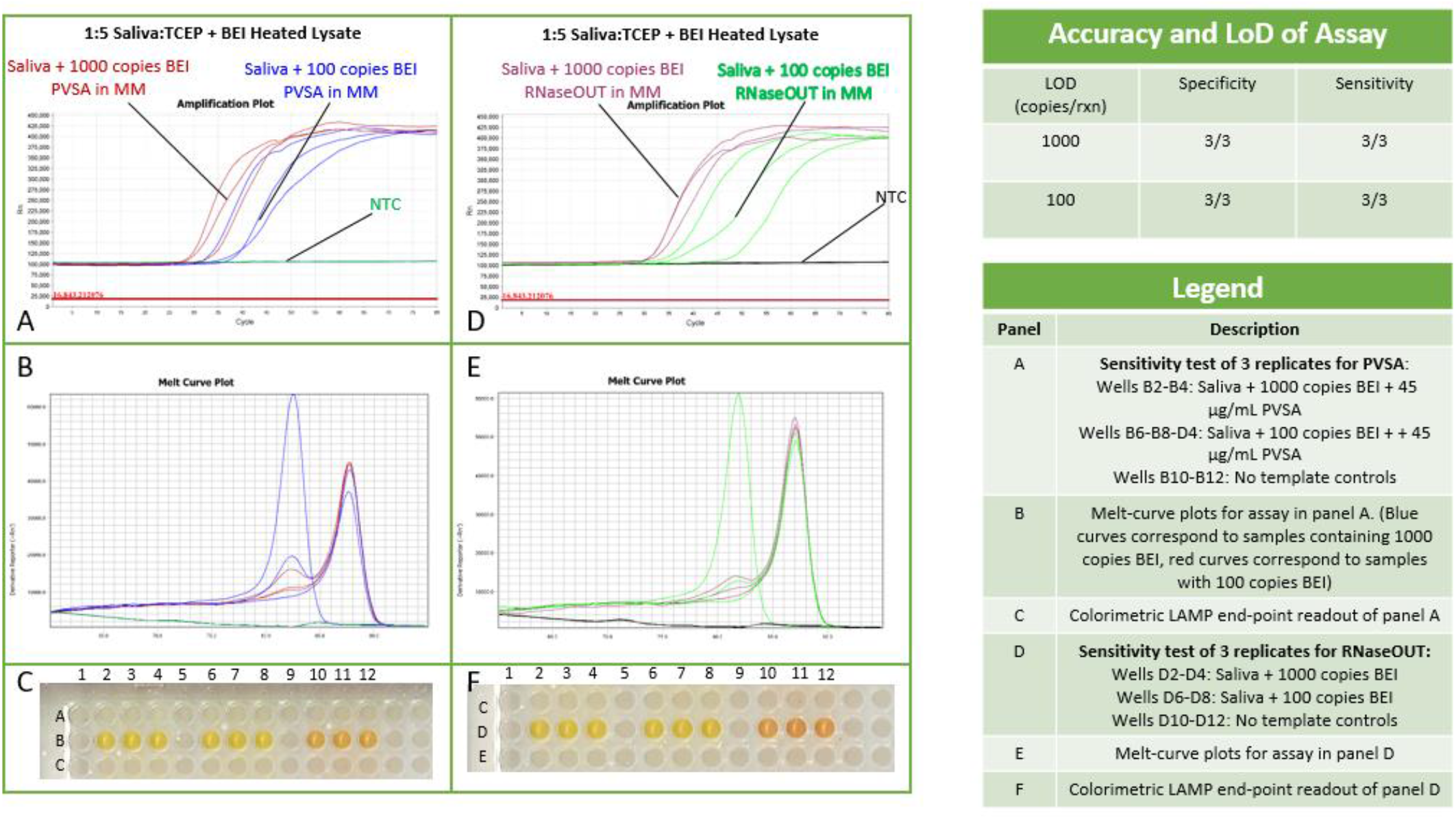
Comparative performance metrics of PVSA vs RNaseOUT

**Figure 5:**
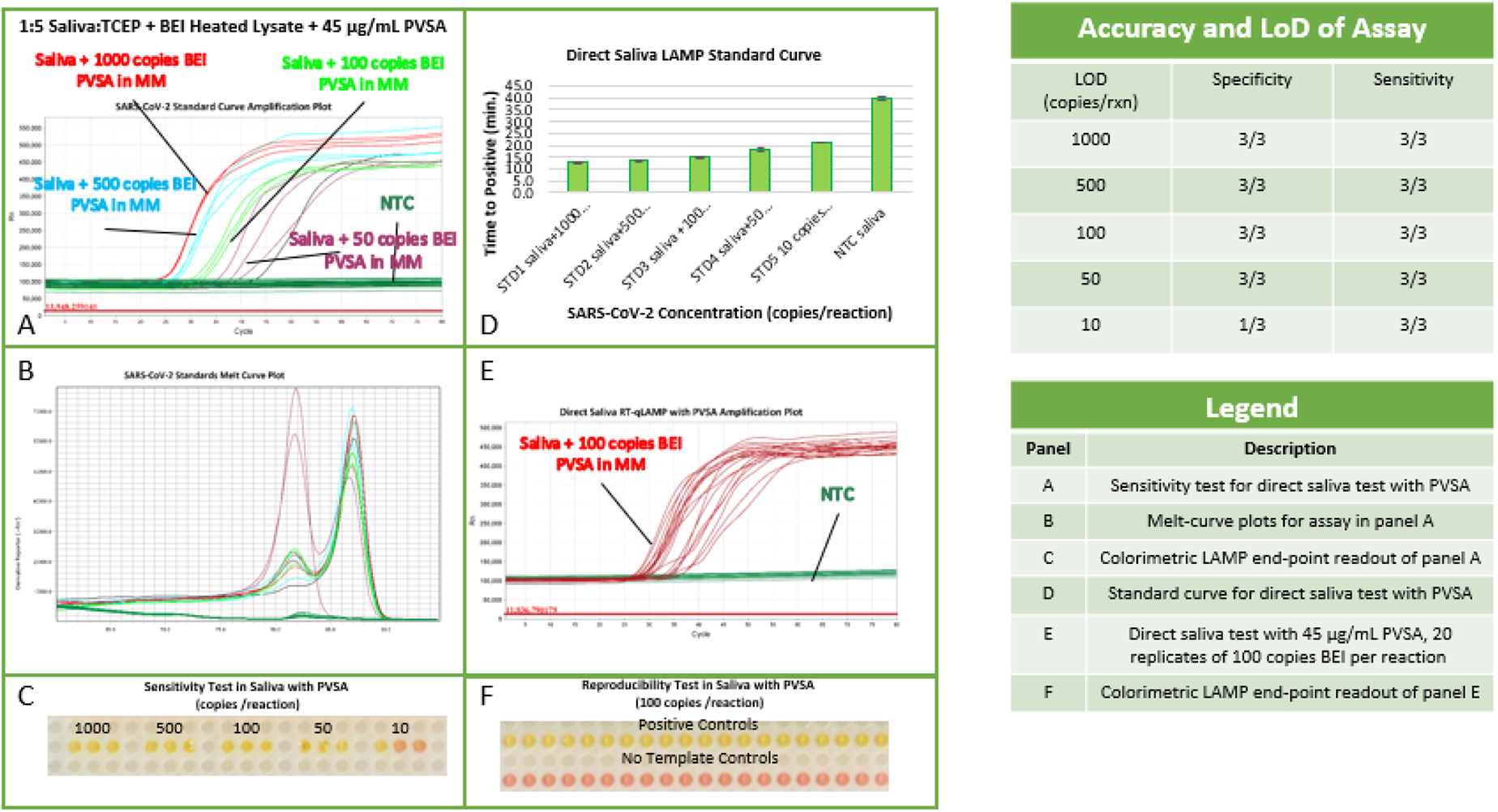
Sensitivity and Specificity testing with PVSA

After establishing that PVSA was at least as effective as RNaseOUT at preserving RNA integrity in saliva, sensitivity tests were conducted to determine the assay’s limit of detection (LoD) with PVSA. Mock saliva samples containing viral concentrations from 1000 copies per reaction to 10 copies per reaction were tested in triplicate. This test showed that the assay retained 100% specificity and sensitivity down to 50 copies per reaction, and 33% specificity and 100% sensitivity at 10 copies per reaction – two-fold lower than with RNaseOUT.

### Stability testing

After optimizing the master mix, stability testing was conducted to determine if pre-plated master mix could withstand prolonged cold storage, thereby enabling scalability and dramatically reducing hands-on time for the assay. Three 384 well plates containing triplicate wells for internal controls, positive controls, no template controls, and tests were prepared. Each plate was frozen at −20°C or −80°C for the following durations: 24 hours, ten days, and one month. At each time point, one plate was removed and thawed at room temperature (Figure 6), then tested by adding and mixing the appropriate sample or control and run on an ABI QuantStudio 12K Flex qPCR instrument with the program described above.

**Figure 6:**
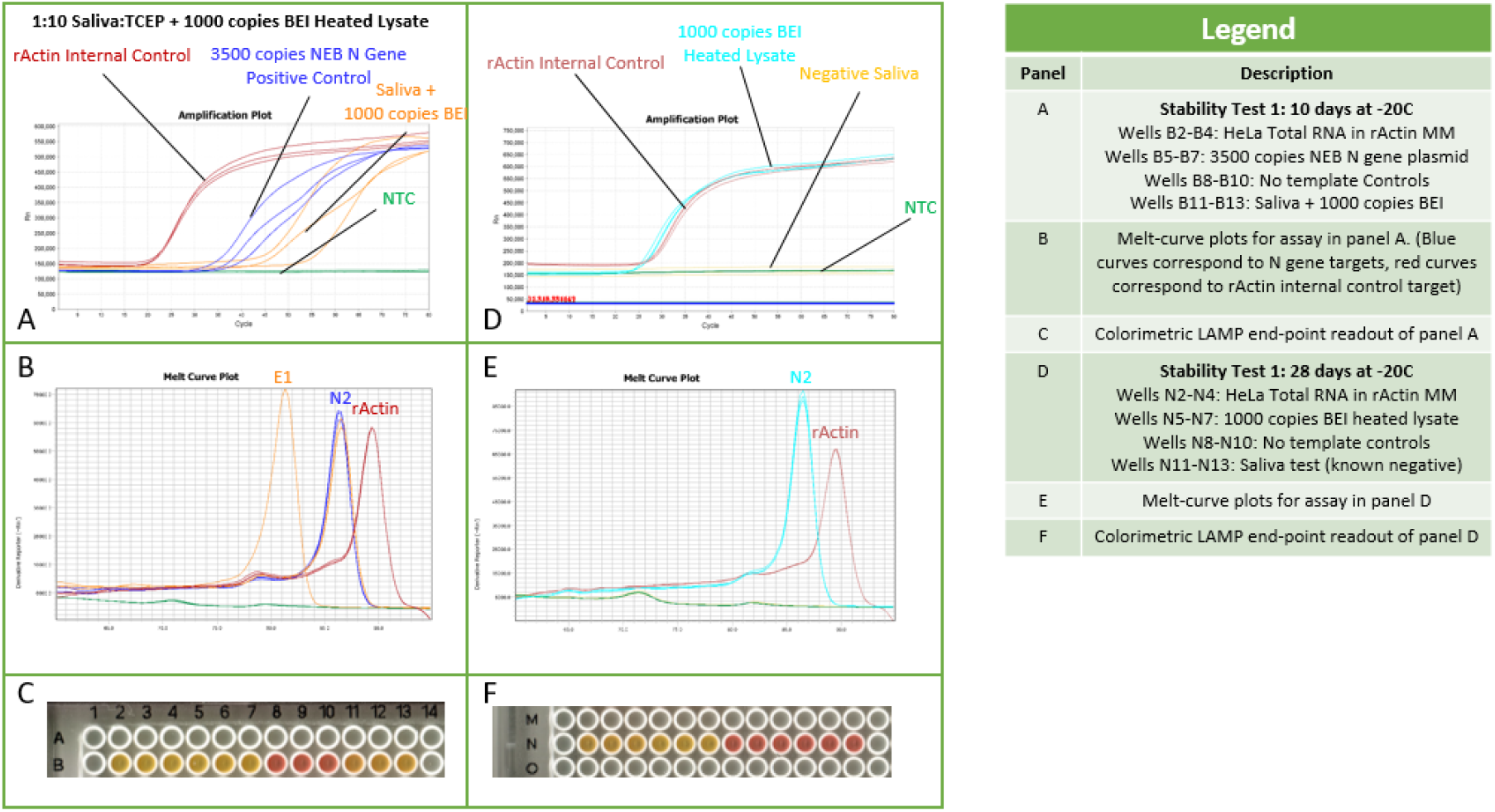
Stability testing

## Discussion

During assay development several components and techniques were identified that proved essential to the reliability of the assay. There were also several short-comings to the colorimetric LAMP assay, and the LAMP method in general was found to have a much steeper learning curve than conventional RT-qPCR. The following factors were critical to the assay’s performance.

### Screening LAMP primers

Thorough in silico screening and bench testing of LAMP primer designs for non-specificity is perhaps the most critical factor in developing a reliable assay, as LAMP reactions typically contain four to 12 primers or more in a reaction mixture. Consequently, non-specific amplification is more probable with LAMP than conventional amplification methods such as PCR, and false positives are a common hazard. Amplification of no template controls were a common occurrence during the early stages of this assay’s development, and inconsistent reproducibility of purified RNA positive controls even with high template concentrations strongly suggested that primer cross reactivity was involved. In this assay, it was found that removing certain loop primers sacrificed some reaction speed but vastly improved specificity and reproducibility. This reliability carried over to direct saliva testing, indicating that the tradeoff in speed for specificity was justified.

Primer specificity testing requires fluorometric validation, to ensure amplification of true LAMP products. This is an essential stage of early RT-LAMP assay development and should not be overlooked. There are several methods now available for monitoring LAMP specificity, including molecular beacons (LAMP-BEAC^27^), fluorescent self-quenching LAMP probes (detection of amplification by release of quenching or DARQ^28^), and quenching of unincorporated amplification signal reporters (QUASR^29^), or nucleic acid specific fluorescent dyes such as SYTO-9. There are advantages of each chemistry, but for this assay SYTO-9 was chosen as the most economic and convenient solution as it was a component already available with the E1700 NEB WarmStart^®^ LAMP kit (DNA &RNA), and as others’ research has shown^30–32^ SYTO-9 is a reliable, non-inhibitory fluorescent reporter in LAMP.

### Reaction additives

Through numerous productive conversations with Nathan Tanner of NEB, and others of the Global LAMP Consortium, several reagents were identified which greatly improved the speed, sensitivity, and reliability of the RT-LAMP assay. Among these key ingredients, guanidine hydrochloride^18^ was found to halve the time to results for the assay, enabling results in 30 minutes or less when two LAMP primer sets were used. It was also found that SYTO-9 fluorescent dye diluted to 0.05X in DMSO improved specificity and sensitivity of the RT-qLAMP assays. This enhancement is consistent with Wang et al. research^33^, that showed 7.5% DMSO could improve LAMP sensitivity and specificity.

Antarctic thermolabile uracil deglycosylase (UDG) was another adjunct suggested by Nathan Tanner, as a means of mitigating carryover contamination and reducing no template amplification. The inclusion of this enzyme was found to be essential to preventing further false positives that plagued early assay development.

Each of these components produced enormous improvements when tested using synthetic or purified RNA extracts. However, when direct saliva tests were conducted the same results were not seen. This inconsistency suggested that RNA template degradation was occurring, and another additive was needed to protect the sample during the assay. As mentioned in the methods, various nuclease inhibitors were tested, but ultimately it was found that a very affordable anionic polymer (PVSA)^20,21^ was the best nuclease inhibitor for RT-LAMP as it improved the limit of detection by two-fold over the best conventional nuclease inhibitor tested, Invitrogen RNaseOUT, at negligible cost (supplementary table 3).

### Sample treatment

Besides the various adjuncts and primer optimizations made to the standard NEB RT-LAMP reaction, it was found that saliva samples required specific treatment to be assayed directly. Several methods for direct saliva RT-LAMP were tested^5,6,22^, some of which included proteinase K for inactivation of nucleases, but it was found that proteinase K remained partially active in the RT-LAMP reaction even after 95°C heat treatment and flash-freezing on dry ice. In our tests, proteinase K often caused false negative results in the RT-qLAMP assays. Residual proteinase K activity was also found to lead to false positive results in no template controls in the colorimetric assays, presumably due to hydrolysis of salivary proteins and other enzymes. For these reasons, proteinase K was omitted from future testing and nuclease inhibitors were focused on instead. It was also determined during assay development that prolonged heat-inactivation used in some protocols severely degraded viral RNA causing false negative results as others have reported^34–37^. Consequently, we chose to use a modified version of Rabe and Cepko’s protocol that involved heat-treating saliva samples at 95°C for five minutes prior to dilution in 2.5 mM TCEP with 1mM EDTA buffer. There are two major advantages to this technique. First, by heat inactivating saliva samples prior to addition of other agents, this dramatically reduces exposure to potential biohazardous material for the tester. As Smyrlaki et al.^21^ confirmed by plaque assay, heat-inactivation at 95°C for 5 minutes is sufficient to completely eliminate SARS-CoV-2 viral activity. The second major advantage of Rabe Cepko’s dilute TCEP solution was that it proved to consistently buffer saliva samples when diluted 5X to 10X, regardless of the collection conditions. Mock tests conducted with saliva samples taken from known negative individuals who drank coffee or even smoked just prior to collection, gave no false positives in any no template controls by colorimetric detection, and all true positive spike-in controls amplified specifically by fluorimetry. This buffer greatly simplified the assay by eliminating the extra time and cost of additional enzymes and processing.

### Future developments

Here several improvements have been suggested for adapting a conventional RT-LAMP kit to be used for high-throughput direct saliva testing of SARS-CoV-2. However, several shortcomings of the assay were found, yet remained unaddressed due to time and other constraints.

Foremost among these were recurrent ambiguous colorimetric results presented by the pH indicator dye used in the colorimetric LAMP mix, phenol red. Phenol red operates within a fairly narrow pH range (6.2 to 8.2^38^) which is convenient for the LAMP reaction, as a mildly buffered solution can demonstrate an observable color change after just a few minutes of amplification. Despite this, a great diversity of colors ranging from cerise, to various shades of orange, to bright yellow can occur during the reaction process. The pH of *any* reaction component will affect these changes in color. Consequently, interpretation of colorimetric results with phenol red indicator can be very subjective. During our tests it was also found that certain additives could even prevent a color change from occurring (e.g. inorganic pyrophosphatase), such that a known positive control sample that showed strong amplification by fluorescence, appeared to remain negative by phenol red colorimetric detection. Had only a colorimetric reporter been used in these reactions these could not have been identified as false negatives.

Fortunately, the current pandemic has spurred on unprecedented innovation in engineering, molecular biology, and chemistry and several affordable alternatives to pH indicator dyes for indirect detection have been found that are adaptable to LAMP assays. These include the use of metal indicator dyes such as hydroxy naphthol blue (HNB) ^3^, eriochrome black T ^39^, and calcein^40^. Numerous methods using dual reporter fluorescent dyes have also been developed over the past few years for both indirect colorimetric end point detection and direct fluorescent measurements, including leuco triphenylmethane dyes^41^, and various fluorescent intercalating dyes^30,32,42^

Another general fault of RT-LAMP assays, including fluorescent assays such as this one, is that these assays are semi-quantitative at best, and so cannot be used to accurately determine viral titer. However, recently developed hue-based LAMP assays such as the open-source smart-phone based eriochrome black T LAMP assay developed by Nguyen et al.^39^ may soon enable point-of-care truly quantitative RT-LAMP for SARS-CoV-2 and other infectious diseases.

Assay cost and shelf-stability are the two remaining hurdles than must be overcome to bring RT-LAMP out of the lab to enable regular affordable testing for all, and these challenges were only partially met by the assay we propose here. Although we succeeded in reducing the cost per test to nearly half that of a conventional kit, $1.80 per test is still far beyond what many people can afford to pay in developing countries. Most of the assay cost came from the proprietary enzymes used in the LAMP reaction mix. This cost could be greatly reduced by substitution of open-source enzymes as Kellner et al.^3^ demonstrated in their rapid beads based SARS-CoV-2 RT-LAMP assay. Also, although we demonstrated that this assay remained stable when frozen for one month, we were unable to conduct shelf-stability tests for longer durations and did not test stability at higher temperatures which is essential for a field deployable assay where refrigeration cannot be guaranteed. Lyophilization of reaction mixtures and stabilization with trehalose sugar can enable room temperature stability of LAMP master mixes^43^, thereby overcoming this obstacle too.Fortunately, as Bektas et al. have shown, with their accessible LAMP-enabled rapid test (ALERT) for SARS-CoV-2^44^, these goals are reasonable and achievable.

## Conclusion

The RT-LAMP assay presented here demonstrates that with some key modifications, a widely available commercial RT-LAMP kit can be adapted for sensitive, robust, timely, and affordable direct detection of SARS-CoV-2 infection from saliva samples. This RT-LAMP formulation, which is stable for at least four weeks at −20°C, provides a low cost, high-throughput method of testing for patient saliva samples directly and can be adapted for future epidemics. SARS-CoV-2 and other infectious diseases will remain a public health burden for the foreseeable future. RT-LAMP assays such as this can help alleviate that burden.

## Data Availability

Any qPCR data or images used in the manuscript are available on request

## Acknowledgements

This work was supported by seed funding through the COVID-19 SUNY Seed Funding Program. We wish to express our special thanks to Nathan Tanner of New England Biolabs for his advice and assistance in troubleshooting during assay development. We also wish to thank Chris Mason of Weill Cornell Medical Research for establishing the Global LAMP Consortium (gLAMP) along with all the other gLAMP members around the world for sharing their expertise, trials and LAMP tribulations without which we could not have gotten where we are.

## Supplementary Information

### Tables

**Suppl. Table 1:** Protocol

| Real-time RT-qLAMP Master Mix (15 uL Reaction) |  |  |  |  |
| --- | --- | --- | --- | --- |
|  |  |  | MM1 (EN*) | MM1 (NEB rActin) |
| Component | Vol (uL) | Final Conc. | For 14 rxns | For 4 rxns |
| CLAMP WarmStart MM (2X) | 7.5 | 1X | 105 | 30 |
| Fluorescent Dye (1x) | 0.80 | 0.05X | 11.20 | 3 |
| LAMP Primer Mix (10X) | 1.5 | 1X | 15 | 6 |
| UDG (1 U/uL) | 0.3 | 0.07 | 4.2 | 1.2 |
| dUTP (100 mM) | 0.1 | 0.7 mM | 1.4 | 0.4 |
| 1.2M GuHCl | 0.5 | 40 mM | 7 | 2 |
| PVSA (900 µg/mL) | 0.75 | 45 µg/mL | 10.5 | 3 |
| Total | 11.45 |  | 139.3 (- Primers) | 39.6 (-Primers) |

1. Combine MM1 components except 10X primers as listed above in 1.5 mL amber tube, then vortex 10 sec at speed 5 to mix, spin down. 2. Transfer 39.6 uL of MM1 to separate amber tube for rActin. 3. Add 15 uL of 10X EN\* primers to first tube and mixed 10 sec by vortexing, quick spun. Add 6 uL of 10X NEB rActin primers to second tube, vortex 10 sec, quick spin down. 4. Add 11.45 uL of MM1 (rActin) to wells B2-B4. Add 11.45 uL of MM1 (E1-LF+N2-LF) to wells B5-B13. 5. Seal plate, centrifuge at 4000 rpm for 5 min., double bag plate in zip lock bags to prevent contamination, then freeze at -20C or -80C until ready to use. 6. Remove plate 20 min. before loading to thaw at RT in darkness. Carefully remove plate seal after thawing, then spiked in 3.55 uL of internal control, positive control, NTC, and sample into triplicate wells, pipetting each well 10X to mix. Reseal plate, centrifuge at 4000 rpm for 5 min. 7. Run plate on QuantStudio 12k Flex or similar real-time qPCR instrument with the following settings: 384 well plate, SYBR, Fast, Relative Std. Curve, 15 uL reaction volume, 10 min. at 25C, 65C for 30 sec X80 cycles with collect data, melt curve.

**Suppl. Table2:**
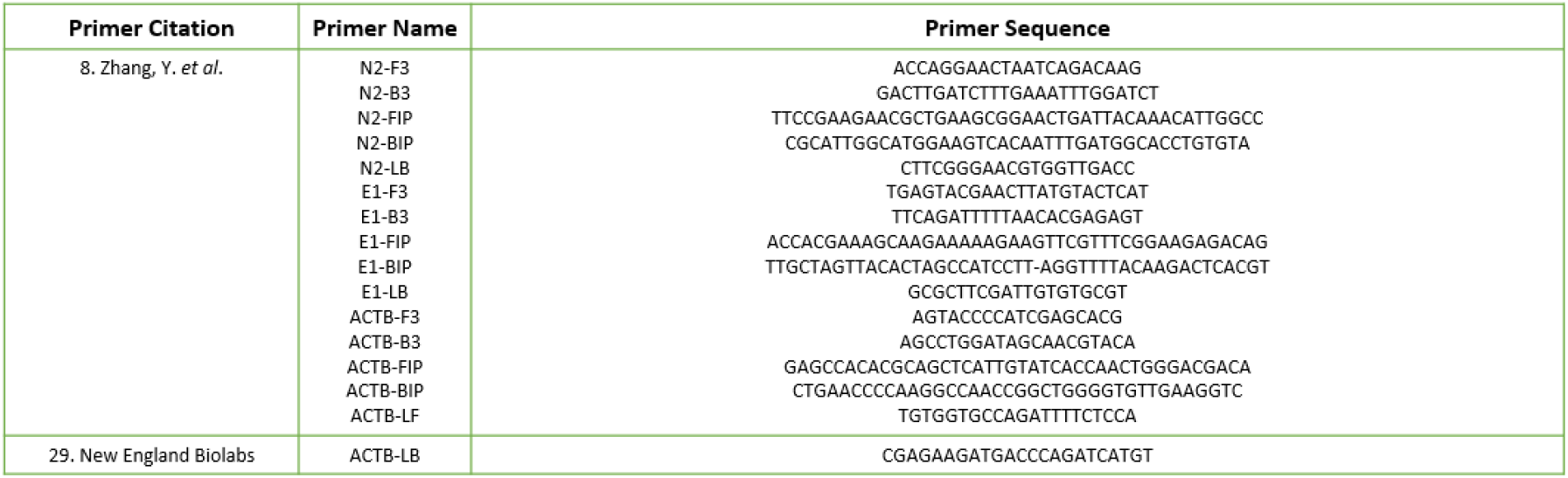
LAMP primer sequences

**Suppl. Table 3.**
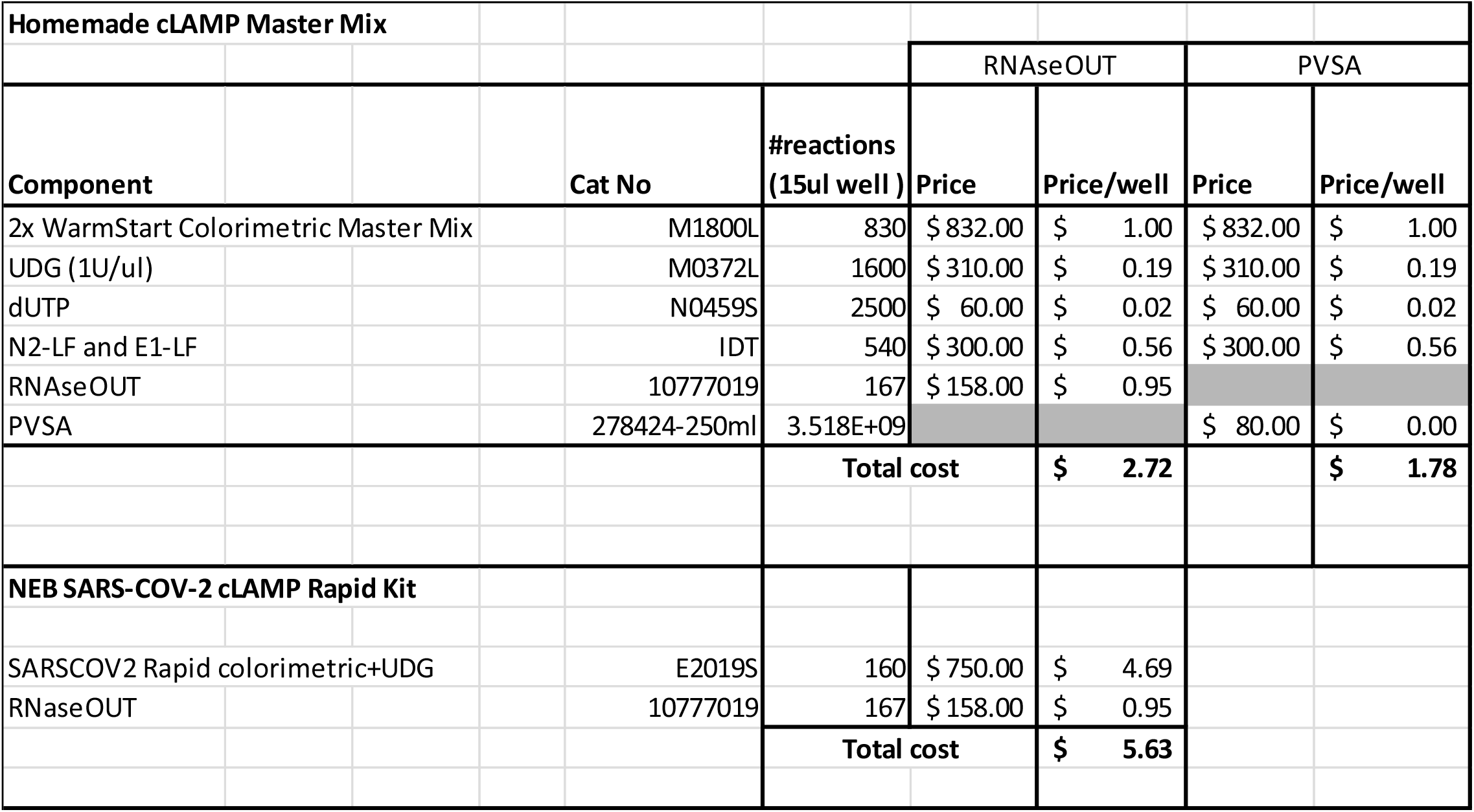
Comparison of cost between RNAseOUT or PVSA based homemade cLAMP mix vs commercial cLAMP Rapid kit

## Notes

### Competing Interest Statement

The authors have declared no competing interest.

### Funding Statement

This project was supported by a seed grant from the State University of New York. This project could not have been completed without the generous donation of colorimetric assay reagents from Nathan Tanner (New England Biolabs) who early on during the pandemic also shared LAMP primer sequences used in this study. This research is part of a global initiative, referred to as gLAMP, to validate LAMP diagnostics for the detection SARS-CoV-2 and we thank all participating members who helped guide our testing protocols from labs across the world.

### Author Declarations

A waiver of ethical approval was received from University at Albany's Institutional Review Board for use of saliva samples in this study

## References

1. Ladha A, Joung J, Abudayyeh O, Gootenberg J, Zhang F. A 5-Min RNA Preparation Method for COVID-19 Detection with RT-QPCR. Infectious Diseases (except HIV/AIDS); 2020. doi:10.1101/2020.05.07.20055947

2. Gray AN, Nichols NM. Facilitating Detection of SARS-CoV-2 Directly from Patient Samples: Precursor Studies with RT-qPCR and Colorimetric RT-LAMP Reagents. :5.

3. Kellner MJ, Ross JJ, Schnabl J, et al. Scalable, rapid and highly sensitive isothermal detection of SARS-CoV-2 for laboratory and home testing. bioRxiv. Published online June 23, 2020:2020.06.23.166397. doi:10.1101/2020.06.23.166397

4. Ikeda M, Imai K, Tabata S, et al. Clinical Evaluation of Self-Collected Saliva by RT-QPCR, Direct RT-QPCR, RT-LAMP, and a Rapid Antigen Test to Diagnose COVID-19. Infectious Diseases (except HIV/AIDS); 2020. doi:10.1101/2020.06.06.20124123

5. Vogels CBF, Watkins AE, Harden CA, et al. SalivaDirect: A simplified and flexible platform to enhance SARS-CoV-2 testing capacity. medRxiv. Published online September 28, 2020:2020.08.03.20167791. doi:10.1101/2020.08.03.20167791

6. Ranoa DRE, Holland RL, Alnaji FG, et al. Saliva-Based Molecular Testing for SARS-CoV-2 that Bypasses RNA Extraction. bioRxiv. Published online June 18, 2020:2020.06.18.159434. doi:10.1101/2020.06.18.159434

7. Kim J-M, Kim HM, Lee EJ, et al. Detection and Isolation of SARS-CoV-2 in Serum, Urine, and Stool Specimens of COVID-19 Patients from the Republic of Korea. Osong Public Health Res Perspect. 2020;11(3):112–117. doi:10.24171/j.phrp.2020.11.3.02

8. Kurtulmus MS, Kazezoglu C, Cakiroglu B, Yilmaz H, Guner AE. The urine foaming test in COVID-19 as a useful tool in diagnosis, prognosis and follow-up: Preliminary results. North Clin Istanb. 2020;7(6):534–540. doi:10.14744/nci.2020.42027

9. El-Tholoth M, Bau HH, Song J. A Single and Two-Stage, Closed-Tube, Molecular Test for the 2019 Novel Coronavirus (COVID-19) at Home, Clinic, and Points of Entry. :21.

10. Dao Thi VL, Herbst K, Boerner K, et al. Screening for SARS-CoV-2 Infections with Colorimetric RT-LAMP and LAMP Sequencing. Infectious Diseases (except HIV/AIDS); 2020. doi:10.1101/2020.05.05.20092288

11. Yang W, Dang X, Wang Q, et al. Rapid and Accurate Detection of Three Genes in SARS-CoV-2 Using RT-LAMP Method. In Review; 2020. doi:10.21203/rs.3.rs-28070/v1

12. Fozouni P, Son S, Díaz de León Derby M, et al. Amplification-free detection of SARS-CoV-2 with CRISPR-Cas13a and mobile phone microscopy. Cell. 2021;184(2):323-333.e9. doi:10.1016/j.cell.2020.12.001

13. Joung J, Ladha A, Saito M, et al. Point-of-Care Testing for COVID-19 Using SHERLOCK Diagnostics. Infectious Diseases (except HIV/AIDS); 2020. doi:10.1101/2020.05.04.20091231

14. Broughton JP, Deng X, Yu G, et al. CRISPR–Cas12-based detection of SARS-CoV-2. Nature Biotechnology. 2020;38(7):870–874. doi:10.1038/s41587-020-0513-4

15. Behrmann O, Bachmann I, Spiegel M, et al. Rapid Detection of SARS-CoV-2 by Low Volume Real-Time Single Tube Reverse Transcription Recombinase Polymerase Amplification Using an Exo Probe with an Internally Linked Quencher (Exo-IQ). Clin Chem. 2020;66(8):1047–1054. doi:10.1093/clinchem/hvaa116

16. Xue G, Li S, Zhang W, et al. Reverse-Transcription Recombinase-Aided Amplification Assay for Rapid Detection of the 2019 Novel Coronavirus (SARS-CoV-2). Anal Chem. 2020;92(14):9699–9705. doi:10.1021/acs.analchem.0c01032

17. Tanner N. Optimized integration of New England Biolabs loop-mediated isothermal amplification (LAMP) reagents with Axxin ISO instruments. :4.

18. Zhang Y, Ren G, Buss J, Barry AJ, Patton GC, Tanner NA. Enhancing Colorimetric LAMP Amplification Speed and Sensitivity with Guanidine Chloride. Biochemistry; 2020. doi:10.1101/2020.06.03.132894

19. Dao Thi VL, Herbst K, Boerner K, et al. A colorimetric RT-LAMP assay and LAMP-sequencing for detecting SARS-CoV-2 RNA in clinical samples. Sci Transl Med. 2020;12(556):eabc7075. doi:10.1126/scitranslmed.abc7075

20. Earl CC, Smith MT, Lease RA, Bundy BC. Polyvinylsulfonic acid: A Low-cost RNase inhibitor for enhanced RNA preservation and cell-free protein translation. Bioengineered. 2017;9(1):90–97. doi:10.1080/21655979.2017.1313648

21. Smyrlaki I, Ekman M, Lentini A, et al. Massive and rapid COVID-19 testing is feasible by extraction-free SARS-CoV-2 RT-PCR. Nature Communications. 2020;11(1):4812. doi:10.1038/s41467-020-18611-5

22. Rabe BA, Cepko C. SARS-CoV-2 detection using isothermal amplification and a rapid, inexpensive protocol for sample inactivation and purification. PNAS. Published online September 7, 2020. doi:10.1073/pnas.2011221117

23. Meagher RJ, Priye A, Light YK, Huang C, Wang E. Impact of Primer Dimers and Self-Amplifying Hairpins on Reverse Transcription Loop-Mediated Isothermal Amplification Detection of Viral RNA. Analyst. 2018;143(8):1924–1933. doi:10.1039/c7an01897e

24. Khorosheva EM, Karymov MA, Selck DA, Ismagilov RF. Lack of correlation between reaction speed and analytical sensitivity in isothermal amplification reveals the value of digital methods for optimization: validation using digital real-time RT-LAMP. Nucleic Acids Res. 2016;44(2):e10–e10. doi:10.1093/nar/gkv877

25. Ding X, Xu Z, Yin K, Sfeir M, Liu C. Dual-Priming Isothermal Amplification (DAMP) for Highly Sensitive and Specific Molecular Detection with Ultralow Nonspecific Signals. Anal Chem. 2019;91(20):12852–12858. doi:10.1021/acs.analchem.9b02582

26. Yu LC, Racevskis J, Webb TE. Regulated Transport of Ribosomal Subunits from Regenerating Rat Liver Nuclei in a Cell-free System. 1972;32:9.

27. Sherrill-Mix S, Hwang Y, Roche AM, et al. LAMP-BEAC: Detection of SARS-CoV-2 RNA Using RT-LAMP and Molecular Beacons. medRxiv. Published online August 19, 2020:2020.08.13.20173757. doi:10.1101/2020.08.13.20173757

28. Tanner NA, Zhang Y, Evans TC. Simultaneous multiple target detection in real-time loop-mediated isothermal amplification. BioTechniques. 2012;53(2):81–89. doi:10.2144/0000113902

29. Quenching of Unincorporated Amplification Signal Reporters in Reverse-Transcription Loop-Mediated Isothermal Amplification Enabling Bright, Single-Step, Closed-Tube, and Multiplexed Detection of RNA Viruses | Analytical Chemistry. Accessed September 27, 2020. https://pubs.acs.org/doi/abs/10.1021/acs.analchem.5b04054

30. Ma B, Yu H, Fang J, Sun C, Zhang M. Employing DNA binding dye to improve detection of Enterocytozoon hepatopenaei in real-time LAMP. Scientific Reports. 2019;9(1):15860. doi:10.1038/s41598-019-52459-0

31. Oscorbin IP, Belousova EA, Zakabunin AI, Boyarskikh UA, Filipenko ML. Comparison of fluorescent intercalating dyes for quantitative loop-mediated isothermal amplification (qLAMP). BioTechniques. 2016;61(1):20–25. doi:10.2144/000114432

32. Seyrig G, Stedtfeld RD, Tourlousse DM, et al. Selection of fluorescent DNA dyes for real-time LAMP with portable and simple optics. J Microbiol Methods. 2015;119:223–227. doi:10.1016/j.mimet.2015.11.004

33. Wang D-G, Brewster JD, Paul M, Tomasula PM. Two Methods for Increased Specificity and Sensitivity in Loop-Mediated Isothermal Amplification. Molecules. 2015;20(4):6048–6059. doi:10.3390/molecules20046048

34. Potential False-Negative Nucleic Acid Testing Results for Severe Acute Respiratory Syndrome Coronavirus 2 from Thermal Inactivation of Samples with Low Viral Loads | Clinical Chemistry | Oxford Academic. Accessed September 27, 2020. https://academic.oup.com/clinchem/article/66/6/794/5815979?rss=1

35. Kampf G, Voss A, Scheithauer S. Inactivation of coronaviruses by heat. J Hosp Infect. Published online March 31, 2020. doi:10.1016/j.jhin.2020.03.025

36. Heat inactivation of serum interferes with the immunoanalysis of antibodies to SARS-CoV-2 | medRxiv. Accessed September 27, 2020. https://www.medrxiv.org/content/10.1101/2020.03.12.20034231v1

37. Evaluation of heating and chemical protocols for inactivating SARS-CoV-2 | bioRxiv. Accessed September 27, 2020. https://www.biorxiv.org/content/10.1101/2020.04.11.036855v1

38. PubChem. Phenol red. Accessed March 21, 2021. https://pubchem.ncbi.nlm.nih.gov/compound/4766

39. Quantification of colorimetric isothermal amplification on the smartphone and its open-source app for point-of-care pathogen detection | Scientific Reports. Accessed March 21, 2021. https://www.nature.com/articles/s41598-020-72095-3

40. Fischbach J, Xander NC, Frohme M, Glökler JF. Shining a light on LAMP assays’ A comparison of LAMP visualization methods including the novel use of berberine. BioTechniques. 2015;58(4):189–194. doi:10.2144/000114275

41. Miyamoto S, Sano S, Takahashi K, Jikihara T. Method for colorimetric detection of double-stranded nucleic acid using leuco triphenylmethane dyes. Analytical Biochemistry. 2015;473:28–33. doi:10.1016/j.ab.2014.12.016

42. Almasi MA, Almasi G. Loop Mediated Isothermal Amplification (LAMP) for Embryo Sex Determination in Pregnant Women at Eight Weeks of Pregnancy. J Reprod Infertil. 2017;18(1):197–204.

43. García-Bernalt Diego J, Fernández-Soto P, Crego-Vicente B, et al. Progress in loop-mediated isothermal amplification assay for detection of Schistosoma mansoni DNA: towards a ready-to-use test. Scientific Reports. 2019;9(1):14744. doi:10.1038/s41598-019-51342-2

44. Bektaş A, Covington MF, Aidelberg G, et al. Accessible LAMP-Enabled Rapid Test (ALERT) for detecting SARS-CoV-2. medRxiv. Published online February 20, 2021:2021.02.18.21251793. doi:10.1101/2021.02.18.21251793

